# Trends in patient attachment to an aging primary care workforce: a population-based serial cross-sectional study in Ontario, Canada

**DOI:** 10.1101/2023.01.19.23284729

**Authors:** Kamila Premji, Michael E Green, Richard H Glazier, Shahriar Khan, Susan E Schultz, Maria Mathews, Steve Nastos, Eliot Frymire, Bridget L Ryan

## Abstract

**Background:** Population aging is a global phenomenon. Resultant healthcare workforce shortages are anticipated. To ensure access to comprehensive primary care, which correlates with improved health outcomes, equity, and costs, data to inform workforce planning are urgently needed.

**Objectives:** To explore temporal trends in early career, mid-career, and near-retirement comprehensive primary care physician characteristics, the medical and social needs of their patients, and the workforce’s capacity to absorb patients of near-retirement physicians. Gender-based workforce trends and trends around alternative practice models were also explored.

**Design:** A serial cross-sectional population-based study using health administrative data.

**Setting:** Ontario, Canada, where most comprehensive primary care is delivered by family physicians (FPs) under universal insurance.

**Participants:** All insured Ontario residents at three time points: 2008 (12,936,360), 2013 (13,447,365), and 2019 (14,388,566) and all Ontario physicians who billed primary care services (2008: 11,566; 2013: 12,693; 2019: 15,054).

**Exposure(s):** Changes in the comprehensive FP workforce over three time periods.

**Main Outcome(s) and Measure(s):** The number and proportion of patients attached to near-retirement comprehensive FPs; the number and proportion of near-retirement comprehensive FPs; the characteristics of patients and their comprehensive FPs.

**Results:** Patient attachment to comprehensive FPs increased over time. The overall FP workforce grew, but the proportion practicing comprehensiveness declined from 77.2% (2008) to 70.7% (2019), with shifts into other/focused scopes of practice across all physician career stages. Over time, an increasing proportion of the comprehensive FP workforce was near retirement age. Correspondingly, an increasing proportion of patients were attached to near-retirement comprehensive FPs. By 2019, 13.9% of comprehensive FPs were 65 years or older, corresponding to 1,695,126 (14.8%) patients. Mean patient age increased, and near-retirement comprehensive FPs served markedly increasing numbers of medically and socially complex patients.

**Conclusions and Relevance:** Primary care is foundational to high-performing health systems, but the sector faces capacity challenges as both patients and physicians age and fewer physicians choose to practice comprehensiveness. Nearly 15% (1.7 million) of Ontarians with a comprehensive FP may lose their physician to retirement by 2025. To serve a growing and increasingly complex patient population, innovative solutions that extend beyond simply growing the FP workforce are needed.

## INTRODUCTION

Primary care is the foundation of high-performing health care systems worldwide,^1^ and can be defined by four core functions (“the 4 Cs”) articulated by Starfield and others: first *Contact* access to the healthcare system, *Continuity* (long-term person-focused care), *Comprehensiveness* (meeting the majority of each patient’s physical and mental health care needs, including prevention, acute care, chronic care, and multimorbidity care), and *Coordination* of care across the healthcare system, including specialty care, hospitals, home care, and community services and support.^1 2^ Access to primary care is associated with improved health outcomes, improved health equity, and reduced health system costs.^3-9^

An essential enabler of primary care access is an adequate health human resource (HHR) supply, but many jurisdictions are grappling with current and impending shortages. For example, 14.5% (4.6 million) Canadians are without a primary care provider.^10^ Virtually every country world-wide is experiencing population aging,^11^ with a high burden of medical complexity^12-15^ and a HHR workforce that is aging into retirement.^16-18^ Concurrently, many countries, including Canada, the United Kingdom, and the United States, are experiencing challenges attracting incoming physicians to primary care as a specialty,^19-22^ and among those who do, a declining proportion are providing primary care reflective of Starfield’s “4 Cs” (hereafter referred to as “comprehensive primary care”); instead, primary care physicians are increasingly limiting their scope of work to subspecialized areas such as sports medicine, dermatology, or palliative care, or to episodic acute care settings, such as walk-in clinics.^23-29^ Moreover, the concentration of women in primary care may further reduce HHR capacity, as women primary care physicians have been found to spend more time with patients^30^ and receive more patient requests outside of appointments than men.^31 32^

In the context of an aging population and shifting workforce demographics, HHR planning requires an understanding of the needs of patients who will soon lose their primary care provider due to retirement, as well as an understanding of the capacity of the remaining and incoming workforce. To anticipate future workforce needs, previous studies often use high-level supply indicators such as number of primary care physicians, and high-level demand indicators such as patient visit rates and durations.^33-36^ In-depth analyses tend to be limited to sub-jurisdictional populations, such as the neighborhood^36^ or early career clinicians,^24^ and do not directly link supply (individual clinicians) to demand (patients served by clinicians).

We conducted an in-depth exploration linking supply and demand at a health system planning level in Ontario, Canada. We examined temporal trends in early career, mid-career, and near-retirement primary care physician characteristics, the medical and social needs of patients attached to these physicians, and the workforce’s capacity to meet the needs of patients of near-retirement physicians. We explored hypothesis-generating differences in gender-based workforce trends, including differences in care provision,^30 31^ and trends around alternative practice models, such as team-based care. As Canadian healthcare planning and delivery are provincial jurisdiction, we focused on the province-level (Ontario). In Ontario, most comprehensive primary care is delivered by family physicians (FPs), most physician services and nearly all residents are covered by government insurance, and health services data are stored centrally in health administrative datasets.

## METHODS

The use of data in this study was authorized under section 45 of Ontario’s Personal Health Information Protection Act (PHIPA) and did not require review by a research ethics board or informed consent. This study followed the Strengthening the Reporting of Observational Studies in Epidemiology (STROBE) reporting guideline.^37^

### Study Design, Population, and Data Sources

We conducted a serial cross-sectional population-level analysis using health administrative data housed at ICES. The study population included all registered Ontario residents covered by the Ontario Health Insurance Plan (OHIP) at three time points: March 31, 2008 (12,936,360), March 31, 2013 (13,447,365), and March 31, 2019 (14,388,566) and all Ontario physicians who billed primary care services (2008: 11,566; 2013: 12,693; 2019: 15,054).

Physician-level and patient-level data came from nine databases which were linked using unique encoded identifiers and analyzed at ICES (Supplement: eMethods).

### Outcomes and Covariates

The primary outcomes were the number and proportion of patients attached to a near-retirement age comprehensive FP over three time points, and the number and proportion of near-retirement age comprehensive FPs over three time points. Based on previous literature finding the average Ontario FP retires at age 70.5 years (with women retiring on average 5 years earlier than men)^38^ and accounting for the time needed to train new physicians,^39^ three different “near-retirement” physician age cut-points were examined: ≥ 55 years, ≥ 65 years, and ≥ 70 years. Comprehensive FPs were defined by applying a previously validated algorithm described below in the Analysis section.^29^

We described the characteristics of both comprehensive FPs and their attached patients over the three time points. Physician characteristics served as exploratory indicators of both supply and, for near-retirement physicians, anticipated demand based on the populations of patients they serve. Patient characteristics served as indicators of demand based on medical and sociodemographic complexity. Detailed data source, cohort, and covariate definitions can be found in the Supplement (eMethods).

### Analysis

For our patient cohort, we created cross-sections of patients attached to comprehensive FPs at three time points: 2008, 2013, 2019.

We began by applying our previously validated algorithm for primary care physician attachment^40^ to the population of OHIP-registered Ontario residents; identifying patients attached to a physician providing longitudinal primary care services based on billing codes and physician-level continuity of care (see Supplement eMethods – continuity of care). We removed patients seen at Community Health Centres because they cannot be attached to a specific physician, patients that the algorithm attached to non-FPs such as pediatricians and surgeons, and patients attached to a FP with missing covariates.

We next created the cohort of FPs linked to the attached patients we identified (2008, 2013, 2019). We stratified our patient and FP cohorts by physician practice type (scope). For this, we used a previously published algorithm for determining comprehensiveness of primary care practice, where physicians are identified as providing comprehensive care if more than half of their services were for core primary care and if these services fell into at least 7 of 22 activity areas.^29^ This resulted in four groups of patients with attachments to four types of FP practice scopes: Comprehensive, Focused (for example, sports medicine or palliative care), Other, and those who worked less than 44 days/year. Focusing on the “comprehensive FP” group, we described the characteristics of these physicians and their patients.

Physician analyses were stratified by physician sex and physician age, including the three “near-retirement” cut-points. Proportions and means with standard deviations were reported for each time point (2008, 2013, 2019).

## RESULTS

### Patient Cohort

Excluding long-term care home residents, the population of OHIP-eligible Ontario residents in the patient cohort over time was 12,863,036 (2008), 13,371,946 (2013), and 14,312,309 (2019), of whom the following were attached to a comprehensive FP: 2008: n = 9,537,353 (77.3%); 2013: n = 10,398,003 (85.1%); 2019: n = 11,480,975 (86.1%) (Figure 1a).

**Figure 1a.**
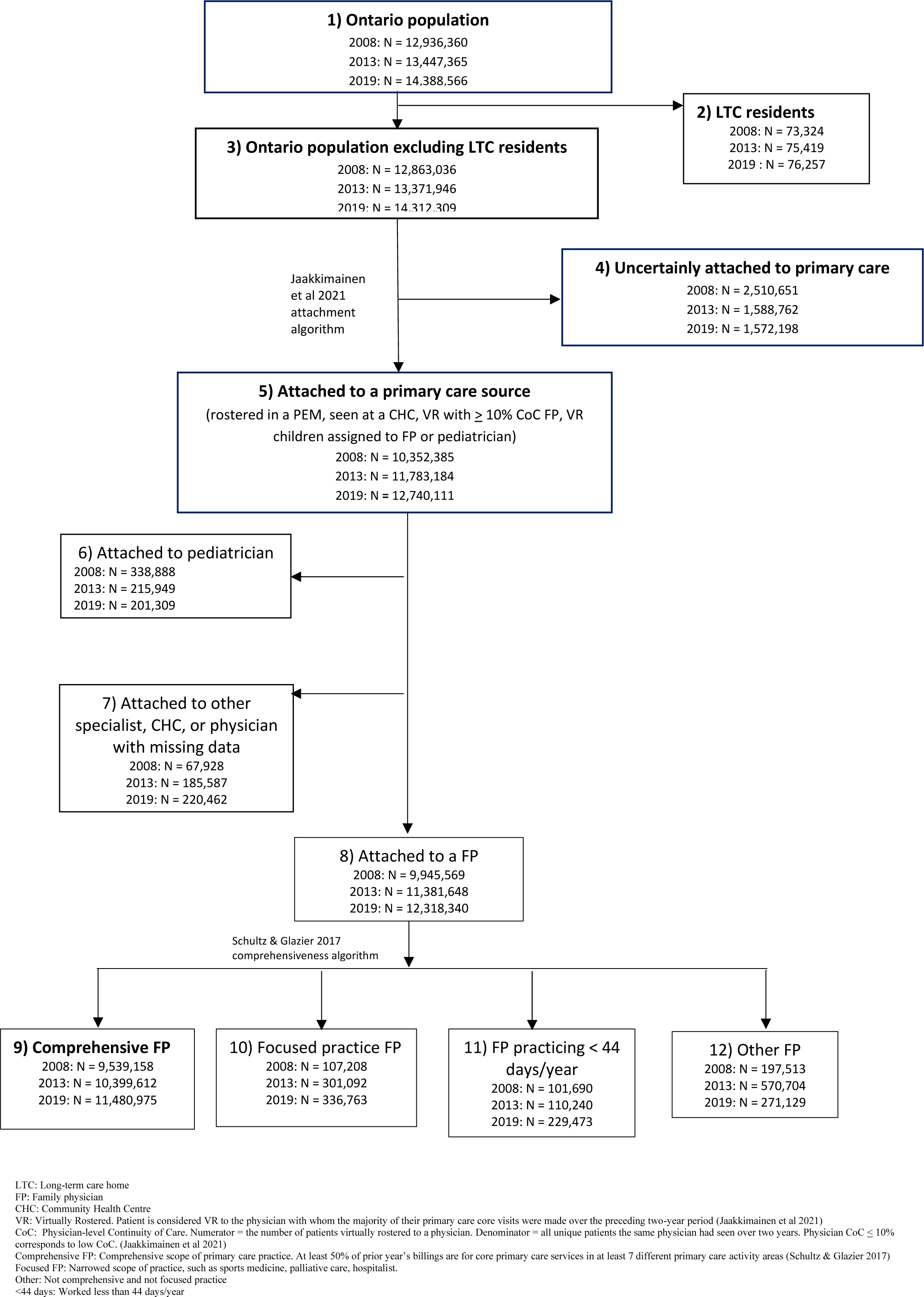
Cohort creation: Patients.

### Physician Cohort

The overall FP workforce grew from 9,944 physicians in 2008 to 13,269 in 2019 (Figure 1b). The proportion of FPs practicing comprehensive primary care declined from 77.2% in 2008 (n = 7,673) to 70.7% in 2019 (n = 9,377) (Supplement: eFigure 1).

**Figure 1b.**
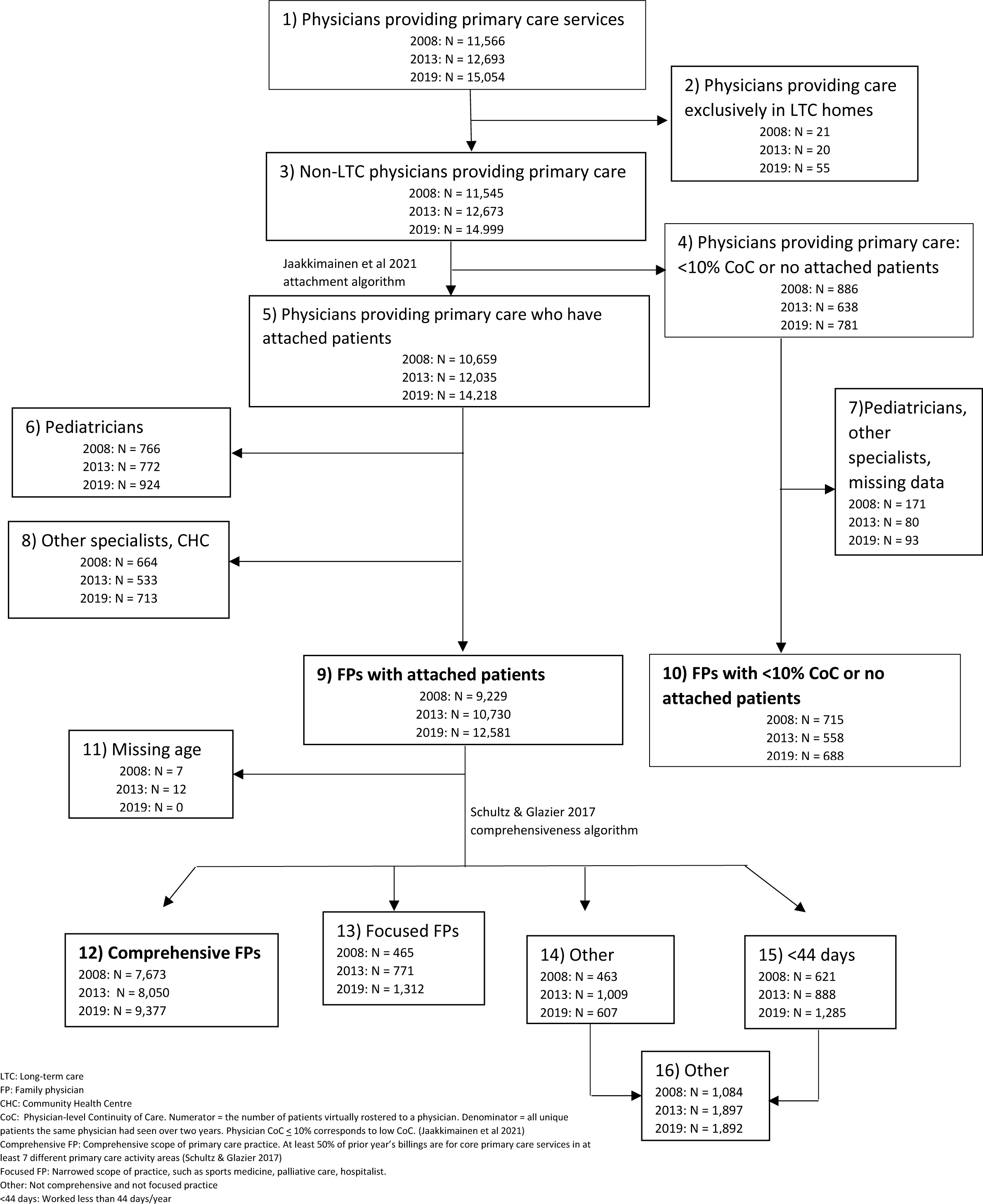
Cohort creation: Physicians.

Table 1 stratifies comprehensive FP data by age and sex. The mean (SD) physician age remained relatively stable over time (2008: 50.3 (11.0) years; 2013: 51.4 (11.8) years; 2019: 49.7 (12.9) years). The mean age (SD) for female physicians was lower than for males at each time point (2008 male 53.0 (10.9) years, female 46.0 (9.7) years; 2013 male 54.7 (11.6) years, female 47.2 (10.6) years; 2019 male 53.1 (13.2) years, female 46.3 (11.6) years). Career stage (years in practice) closely followed physician age group for both males and females, and the youngest cohort (age <35) comprised an increasing proportion of the workforce over time, shifting from 7.7% in 2008 to 15.1% in 2019. The older cohorts were also found to comprise an increasing proportion of the workforce over time, and the absolute numbers of older physicians increased.

**Table 1.** Practice characteristics of comprehensive family physicians

|  |  | <35 Years |  |  | 35-44 Years |  |  | 45-54 Years |  |  | 55-64 Years |  |  | 65-69 Years |  |  | 70+ Years |  |  | Total Comprehensive FPs |  |  |
| --- | --- | --- | --- | --- | --- | --- | --- | --- | --- | --- | --- | --- | --- | --- | --- | --- | --- | --- | --- | --- | --- | --- |
|  |  | Total | M | F | Total | M | F | Total | M | F | Total | M | F | Total | M | F |  | M | F | Total | M | F |
| Comp. FPs<br>N (%) | 2008 | 592<br>(7.7) | 211<br>(35.6) | 381<br>(64.4) | 1877<br>(24.5) | 922<br>(49.1) | 955<br>(50.9) | 2467<br>(32.2) | 1422<br>(57.6) | 1045<br>(42.4) | 1972<br>(25.7) | 1522<br>(77.2) | 450<br>(22.8) | 409<br>(5.3) | 347<br>(84.8) | 62<br>(15.2) | 356<br>(4.6) | 319<br>(89.6) | 37<br>(10.4) | 7673<br>(100.0) | 4743<br>(61.8) | 2930<br>(38.2) |
|  | 2013 | 741<br>(9.2) | 245<br>(33.1) | 496<br>(66.9) | 1666<br>(20.7) | 674<br>(40.5) | 992<br>(59.5) | 2312<br>(28.7) | 1227<br>(53.1) | 1085<br>(46.9) | 2170<br>(27.0) | 1415<br>(65.2) | 755<br>(34.8) | 707<br>(8.8) | 576<br>(81.5) | 131<br>(18.5) | 454<br>(5.6) | 392<br>(86.3) | 62<br>(13.7) | 8050<br>(100.0) | 4529<br>(56.3) | 3521<br>(43.7) |
|  | 2019 | 1414<br>(15.1) | 528<br>(37.3) | 886<br>(62.7) | 2135<br>(22.8) | 806<br>(37.8) | 1329<br>(62.2) | 2242<br>(23.9) | 1048<br>(46.7) | 1194<br>(53.3) | 2279<br>(24.3) | 1290<br>(56.6) | 989<br>(43.4) | 708<br>(7.6) | 519<br>(73.3) | 189<br>(26.7) | 599<br>(6.4) | 505<br>(84.3) | 94<br>(15.7) | 9377<br>(100.0) | 4696<br>(50.1) | 4681<br>(49.9) |
| Years in<br>pract.<br>(mean<br>(SD)) | 2008 | 6.0<br>(±2.3) | 6.3<br>(±2.3) | 5.9<br>(±2.2) | 14.4<br>(±3.9) | 14.7<br>(±3.8) | 14.1<br>(±3.9) | 23.7<br>(±4.2) | 23.8<br>(±4.2) | 23.5<br>(±4.2) | 33.4<br>(±4.4) | 33.6<br>(±4.2) | 32.8<br>(±4.8) | 41.3<br>(±3.0) | 41.2<br>(±3.0) | 42.0<br>(±3.2) | 48.0<br>(±5.1) | 48.0<br>(±4.9) | 47.8<br>(±6.4) | 24.6<br>(±11.4) | 27.3<br>(±11.2) | 20.2<br>(±10.1) |
|  | 2013 | 5.7<br>(±2.1) | 5.4<br>(±2.1) | 5.9<br>(±2.1) | 13.8<br>(±4.2) | 14.0<br>(±4.2) | 13.7<br>(±4.1) | 23.9<br>(±4.2) | 23.9<br>(±4.0) | 23.8<br>(±4.4) | 33.2<br>(±4.4) | 33.6<br>(±4.4) | 32.5<br>(±4.5) | 41.2<br>(±3.5) | 41.1<br>(±3.4) | 41.6<br>(±4.0) | 48.7<br>(±4.9) | 48.7<br>(±4.9) | 49.0<br>(±4.9) | 25.6<br>(±12.3) | 28.8<br>(±12.1) | 21.4<br>(±11.1) |
|  | 2019 | 5.8<br>(±2.0) | 5.7<br>(±2.0) | 5.8<br>(±1.9) | 12.5<br>(±4.2) | 12.5<br>(±4.4) | 12.5<br>(±4.0) | 23.7<br>(±4.7) | 23.9<br>(±4.7) | 23.5<br>(±4.6) | 33.3<br>(±4.7) | 33.4<br>(±4.5) | 33.2<br>(±4.9) | 40.8<br>(±3.6) | 41.0<br>(±3.4) | 40.3<br>(±4.0) | 48.5<br>(±5.1) | 48.4<br>(±5.3) | 48.7<br>(±4.1) | 23.7<br>(±13.4) | 27.0<br>(±13.8) | 20.3<br>(±12.0) |
| Roster<br>size<br>(mean<br>(SD)) | 2008 | 638.3<br>(±622.5<br>) | 790.7<br>(±722.0<br>) | 553.9<br>(±542.7<br>) | 1131.8<br>(±873.2<br>) | 1323.5<br>(±981.3<br>) | 946.7<br>(±707.0<br>) | 1345.1<br>(±920.7<br>) | 1470.3<br>(±996.7<br>) | 1174.6<br>(±774.4<br>) | 1432.1<br>(±945.2<br>) | 1494.0<br>(±961.5<br>) | 1222.7<br>(±856.4<br>) | 1123.1<br>(±955.5<br>) | 1186.1<br>(±981.7<br>) | 770.7<br>(±701.1<br>) | 566.3<br>(±770.9<br>) | 584.9<br>(±785.4<br>) | 406.5<br>(±618.7<br>) | 1212.8<br>(±927.0<br>) | 1338.8<br>(±991.1<br>) | 1008.8<br>(±770.0<br>) |
|  | 2013 | 620.0<br>(±605.9<br>) | 725.2<br>(±690.9<br>) | 568.0<br>(±552.6<br>) | 1152.8<br>(±836.0<br>) | 1348.6<br>(±935.1<br>) | 1019.7<br>(±732.6<br>) | 1407.1<br>(±927.1<br>) | 1567.8<br>(±1013.4<br>) | 1225.4<br>(±780.2<br>) | 1490.2<br>(±894.6<br>) | 1593.1<br>(±937.6<br>) | 1297.2<br>(±772.4<br>) | 1366.1<br>(±905.8<br>) | 1420.3<br>(±921.3<br>) | 1128.0<br>(±794.3<br>) | 898.1<br>(±895.7<br>) | 946.7<br>(±922.9<br>) | 591.1<br>(±622.7<br>) | 1272.1<br>(±909.2<br>) | 1425.0<br>(±975.2<br>) | 1075.4<br>(±773.4<br>) |
|  | 2019 | 734.0<br>(±644.2<br>) | 834.7<br>(±712.0<br>) | 674.0<br>(±592.4<br>) | 1074.5<br>(±720.3<br>) | 1217.2<br>(±841.6<br>) | 987.9<br>(±620.1<br>) | 1394.8<br>(±876.2<br>) | 1529.3<br>(±946.5<br>) | 1276.7<br>(±791.2<br>) | 1405.6<br>(±847.2<br>) | 1531.6<br>(±902.2<br>) | 1241.1<br>(±738.3<br>) | 1434.4<br>(±900.5<br>) | 1502.5<br>(±932.8<br>) | 1247.6<br>(±777.3<br>) | 1098.0<br>(±804.3<br>) | 1125.7<br>(±815.1<br>) | 949.2<br>(±729.6<br>) | 1208.9<br>(±837.4<br>) | 1351.9<br>(±908.8<br>) | 1065.4<br>(±731.6<br>) |
| Core PC<br>visits<br>(mean<br>(SD)) | 2008 | 6.2<br>(±2.7) | 6.2<br>(±2.8) | 6.2<br>(±2.7) | 7.3<br>(±4.2) | 7.5<br>(±5.6) | 7.2<br>(±2.3) | 7.3<br>(±2.3) | 7.4<br>(±2.5) | 7.3<br>(±2.1) | 7.7<br>(±2.6) | 7.7<br>(±2.6) | 7.7<br>(±2.4) | 7.5<br>(±3.1) | 7.6<br>(±3.2) | 6.9<br>(±2.7) | 6.8<br>(±3.5) | 6.9<br>(±3.5) | 6.2<br>(±2.9) | 7.3<br>(±3.1) | 7.4<br>(±3.5) | 7.1<br>(±2.4) |
|  | 2013 | 5.3<br>(±2.3) | 5.4<br>(±2.3) | 5.3<br>(±2.3) | 6.3<br>(±2.1) | 6.2<br>(±2.2) | 6.3<br>(±2.0) | 6.5<br>(±2.4) | 6.6<br>(±2.7) | 6.4<br>(±2.0) | 6.7<br>(±2.8) | 6.8<br>(±3.2) | 6.4<br>(±1.9) | 6.9<br>(±2.4) | 6.9<br>(±2.4) | 7.0<br>(±2.3) | 7.3<br>(±4.0) | 7.5<br>(±4.2) | 6.5<br>(±2.4) | 6.5<br>(±2.6) | 6.6<br>(±2.9) | 6.3<br>(±2.1) |
|  | 2019 | 5.6<br>(±2.5) | 5.5<br>(±2.6) | 5.6<br>(±2.4) | 6.0<br>(±2.5) | 5.9<br>(±2.8) | 6.0<br>(±2.4) | 6.1<br>(±2.1) | 6.1<br>(±2.3) | 6.1<br>(±1.9) | 6.1<br>(±2.1) | 6.2<br>(±2.3) | 6.0<br>(±1.8) | 6.4<br>(±2.2) | 6.5<br>(±2.3) | 6.2<br>(±2.0) | 6.7<br>(±3.0) | 6.5<br>(±2.9) | 7.2<br>(±3.1) | 6.0<br>(±2.3) | 6.1<br>(±2.5) | 6.0<br>(±2.2) |
| Pt age<br>(mean<br>(SD)) | 2008 | 27.9<br>(±13.8) | 29.4<br>(±14.0) | 27.1<br>(±13.6) | 31.7<br>(±11.7) | 32.8<br>(±12.6) | 30.5<br>(±10.7) | 34.3<br>(±11.9) | 35.4<br>(±12.5) | 32.7<br>(±10.8) | 36.7<br>(±13.1) | 37.6<br>(±13.2) | 33.7<br>(±12.2) | 35.1<br>(±16.2) | 36.0<br>(±16.1) | 30.5<br>(±15.9) | 28.2<br>(±18.5) | 28.5<br>(±18.5) | 25.5<br>(±17.8) | 33.5<br>(±13.2) | 34.9<br>(±13.8) | 31.3<br>(±11.8) |
|  | 2013 | 28.2<br>(±13.7) | 30.0<br>(±13.7) | 27.4<br>(±13.6) | 34.0<br>(±10.8) | 35.0<br>(±11.6) | 33.4<br>(±10.1) | 36.4<br>(±10.7) | 37.8<br>(±11.2) | 34.8<br>(±9.9) | 39.4<br>(±10.7) | 40.5<br>(±11.1) | 37.3<br>(±9.8) | 40.9<br>(±12.6) | 42.0<br>(±12.4) | 36.3<br>(±12.7) | 39.1<br>(±17.0) | 39.7<br>(±17.1) | 35.0<br>(±16.0) | 36.5<br>(±12.1) | 38.5<br>(±12.5) | 34.0<br>(±11.2) |
|  | 2019 | 31.8<br>(±14.5) | 33.5<br>(±14.2) | 30.7<br>(±14.5) | 36.4<br>(±10.9) | 37.1<br>(±11.8) | 36.0<br>(±10.3) | 38.4<br>(±9.8) | 39.4<br>(±10.6) | 37.5<br>(±9.0) | 40.6<br>(±10.5) | 42.0<br>(±10.8) | 38.7<br>(±9.8) | 43.0<br>(±11.5) | 43.9<br>(±11.6) | 40.8<br>(±10.9) | 43.3<br>(±14.3) | 43.6<br>(±14.5) | 41.2<br>(±13.1) | 38.1<br>(±12.0) | 40.0<br>(±12.3) | 36.2<br>(±11.3) |
| Prop. Fem.<br>Pts<br>(mean<br>(SD)) | 2008 | 55.7<br>(±15.1) | 46.9<br>(±10.7) | 60.7<br>(±14.9) | 55.2<br>(±13.2) | 46.2<br>(±7.5) | 63.8<br>(±11.6) | 54.3<br>(±13.0) | 46.3<br>(±7.4) | 65.3<br>(±10.9) | 51.0<br>(±11.0) | 46.8<br>(±7.0) | 65.0<br>(±10.7) | 49.5<br>(±11.1) | 47.3<br>(±8.5) | 61.5<br>(±15.7) | 47.8<br>(±13.2) | 46.7<br>(±11.1) | 57.6<br>(±22.6) | 53.2<br>(±12.9) | 46.6<br>(±7.8) | 64.0<br>(±12.1) |
|  | 2013 | 55.3<br>(±15.6) | 47.8<br>(±13.7) | 59.0<br>(±15.1) | 55.1<br>(±12.1) | 46.1<br>(±8.3) | 61.2<br>(±10.4) | 53.7<br>(±12.3) | 45.6<br>(±7.4) | 62.9<br>(±9.9) | 52.4<br>(±12.1) | 45.9<br>(±7.5) | 64.7<br>(±9.3) | 48.9<br>(±10.1) | 45.9<br>(±7.2) | 62.2<br>(±10.5) | 49.6<br>(±12.2) | 47.2<br>(±10.4) | 64.8<br>(±11.9) | 53.1<br>(±12.5) | 46.1<br>(±8.3) | 62.3<br>(±11.0) |
|  | 2019 | 54.3<br>(±13.7) | 47.7<br>(±11.2) | 58.2<br>(±13.6) | 54.3<br>(±11.8) | 45.0<br>(±8.2) | 59.9<br>(±10.0) | 53.5<br>(±11.2) | 45.4<br>(±7.6) | 60.6<br>(±8.9) | 52.4<br>(±11.8) | 44.8<br>(±7.8) | 62.2<br>(±8.5) | 49.9<br>(±11.7) | 45.1<br>(±7.9) | 63.0<br>(±10.2) | 48.2<br>(±9.9) | 45.9<br>(±8.1) | 60.7<br>(±9.6) | 52.9<br>(±12.0) | 45.5<br>(±8.4) | 60.4<br>(±10.3) |
| FTE<br>(N (%)) | 2008 | 290<br>(49.0) | 146<br>(50.3) | 144<br>(49.7) | 1210<br>(64.5) | 754<br>(62.3) | 456<br>(37.7) | 1802<br>(73.0) | 1173<br>(65.1) | 629<br>(34.9) | 1481<br>(75.1) | 1209<br>(81.6) | 272<br>(18.4) | 239<br>(58.4) | 220<br>(92.1) | 19<br>(8.0) | 114<br>(32.0) | 107<br>(93.9) | 7<br>(6.1) | 5136<br>(66.9) | 3609<br>(70.3) | 1527<br>(29.7) |
|  | 2013 | 335<br>(45.4) | 152<br>(45.4) | 183<br>(54.6) | 1073<br>(64.4) | 556<br>(51.8) | 517<br>(48.2) | 1694<br>(73.3) | 1014<br>(59.9) | 680<br>(40.1) | 1634<br>(75.3) | 1156<br>(70.8) | 478<br>(29.3) | 474<br>(67.0) | 415<br>(87.6) | 59<br>(12.5) | 189<br>(41.6) | 177<br>(93.7) | 12<br>(6.4) | 5399<br>(67.1) | 3470<br>(64.3) | 1929<br>(35.7) |
|  | 2019 | 734<br>(51.9) | 351<br>(47.8) | 383<br>(52.2) | 1401<br>(65.6) | 628<br>(44.8) | 773<br>(55.2) | 1722<br>(76.8) | 881<br>(51.2) | 841<br>(48.8) | 1681<br>(73.8) | 1052<br>(62.6) | 629<br>(37.4) | 514<br>(72.6) | 402<br>(78.2) | 112<br>(21.8) | 327<br>(54.6) | 288<br>(88.1) | 39<br>(11.9) | 6379<br>(68.0) | 3602<br>(56.5) | 2777<br>(43.5) |

Comp. FPs: Comprehensive family physicians; Pract.: Practice; PC: Primary care; Pt(s): Patient(s); Prop: Proportion; Fem: Female; FTE: Full-time equivalent

Among family physicians with patient attachments, a shift away from comprehensiveness and into other/focused scopes of practice was seen across all physician age groups, with the most pronounced shifts in the youngest and oldest physician groups (Supplement: eTable 1). Instead of comprehensive primary care, these FPs increasingly worked in focused or other scopes of practice. The proportion of FPs identified as practicing exclusively without patient attachments or in low-continuity (“walk-in clinic”) settings fluctuated: 2008: 7.2% (n = 715), 2013: 4.9% (n = 558); 2019: 5.2% (n = 688) (Figure 1b).

### Temporal Trends of Near-Retirement Comprehensive Family Physicians and their Patients

When looking at our three near-retirement cut-points (55+, 65+, 70+) over time, an increasing proportion of the comprehensive FP workforce was near retirement age (Figure 2).

**Figure 2.**
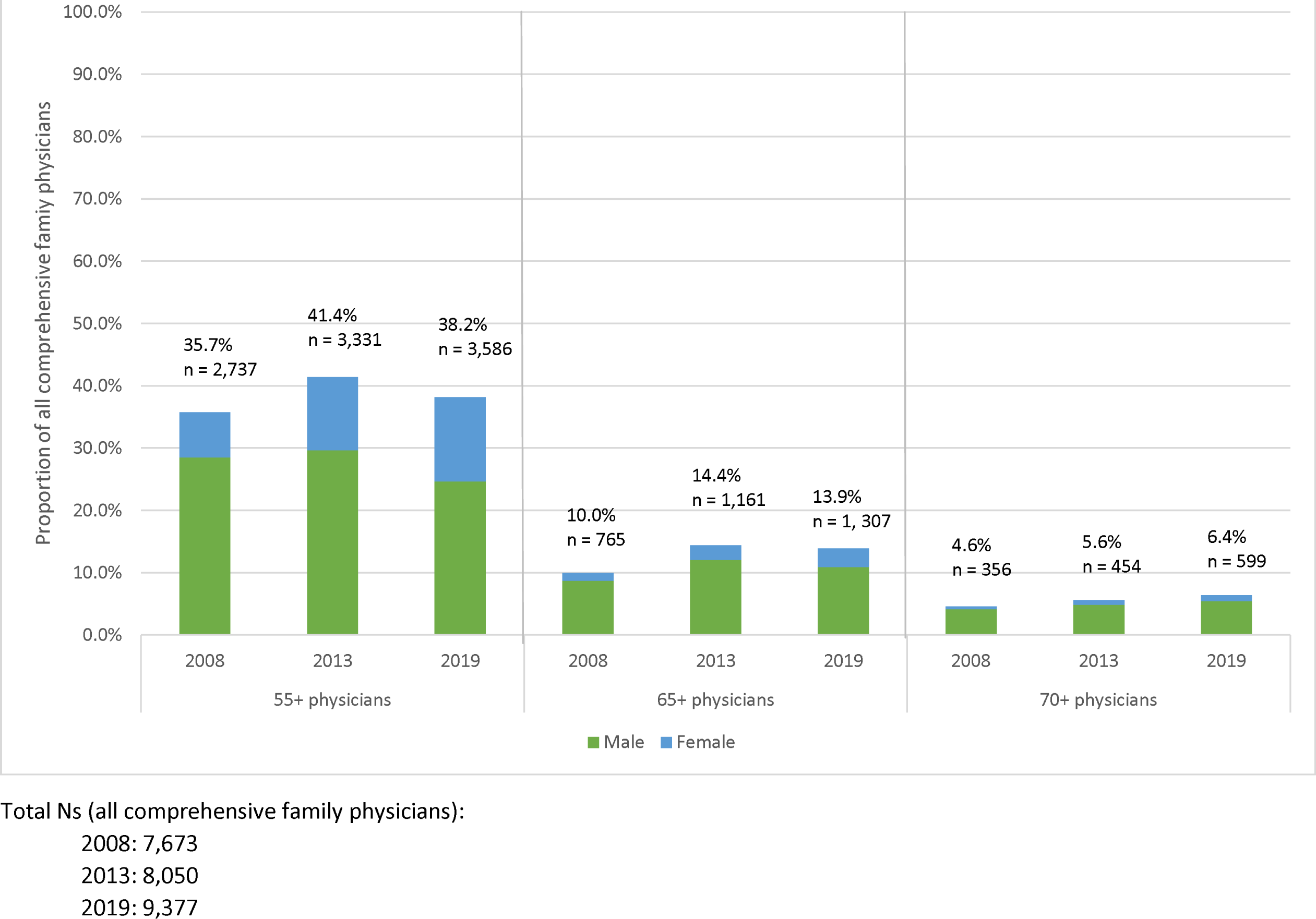
Comprehensive family physicians by near-retirement group, year, and sex.

Correspondingly, an increasing proportion of patients were attached to near-retirement comprehensive FPs (Table 2). In the 55+ age group, the proportion of comprehensive FPs increased from 35.7% in 2008 to 38.2% in 2019. In 2019, this corresponded to 3,586 physicians and 4,935,992 (43.0%) patients (2019). In the 65+ group, the proportion increased from 10.0% in 2008 to 13.9% in 2019 (1,307 physicians, 1,695,126 (14.8%) patients). In the 70+ age group, the proportion increased from 4.6% in 2008 to 6.4% in 2019 (599 physicians, 666,000 (5.8%) patients).

**Table 2.**
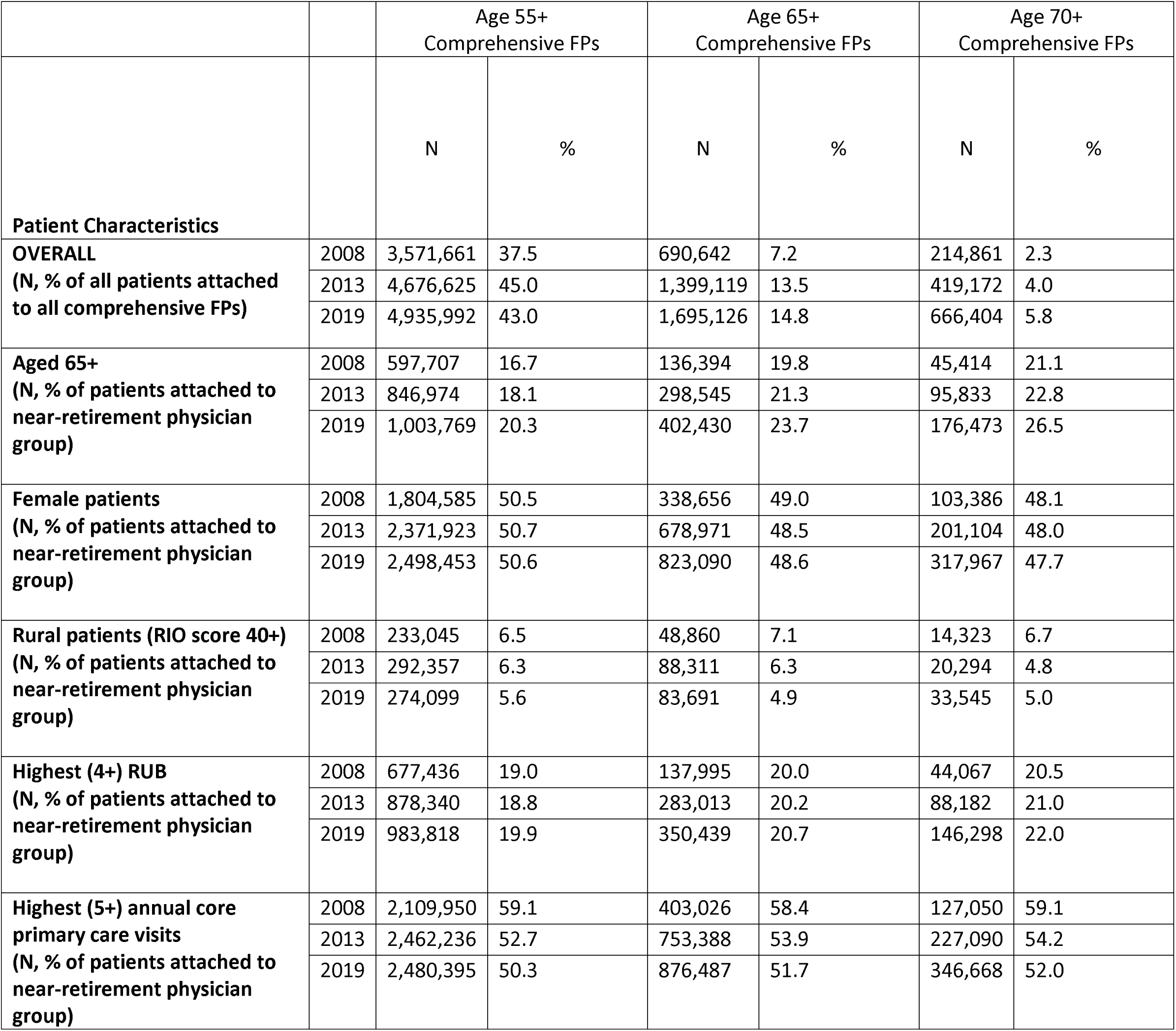
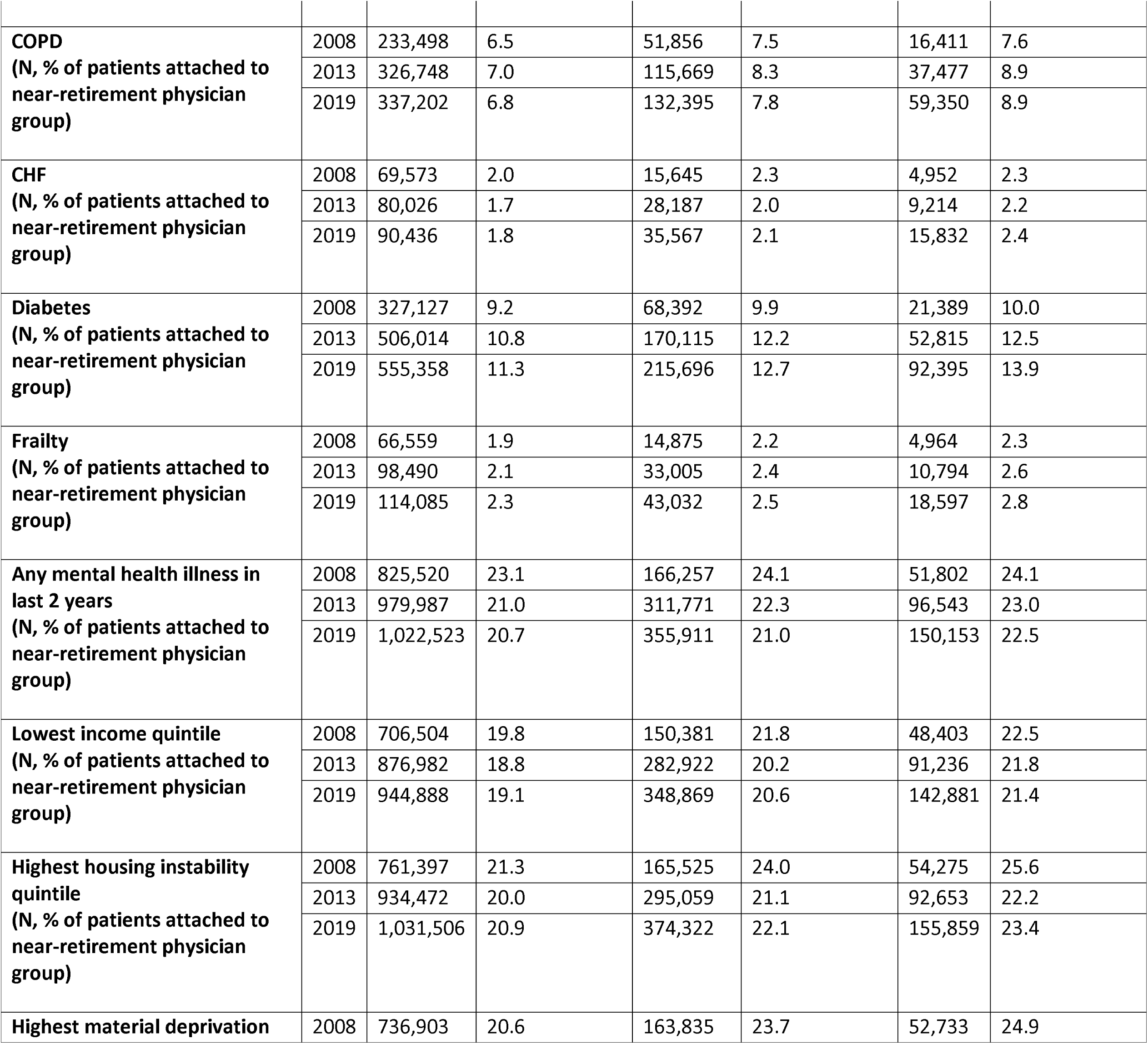
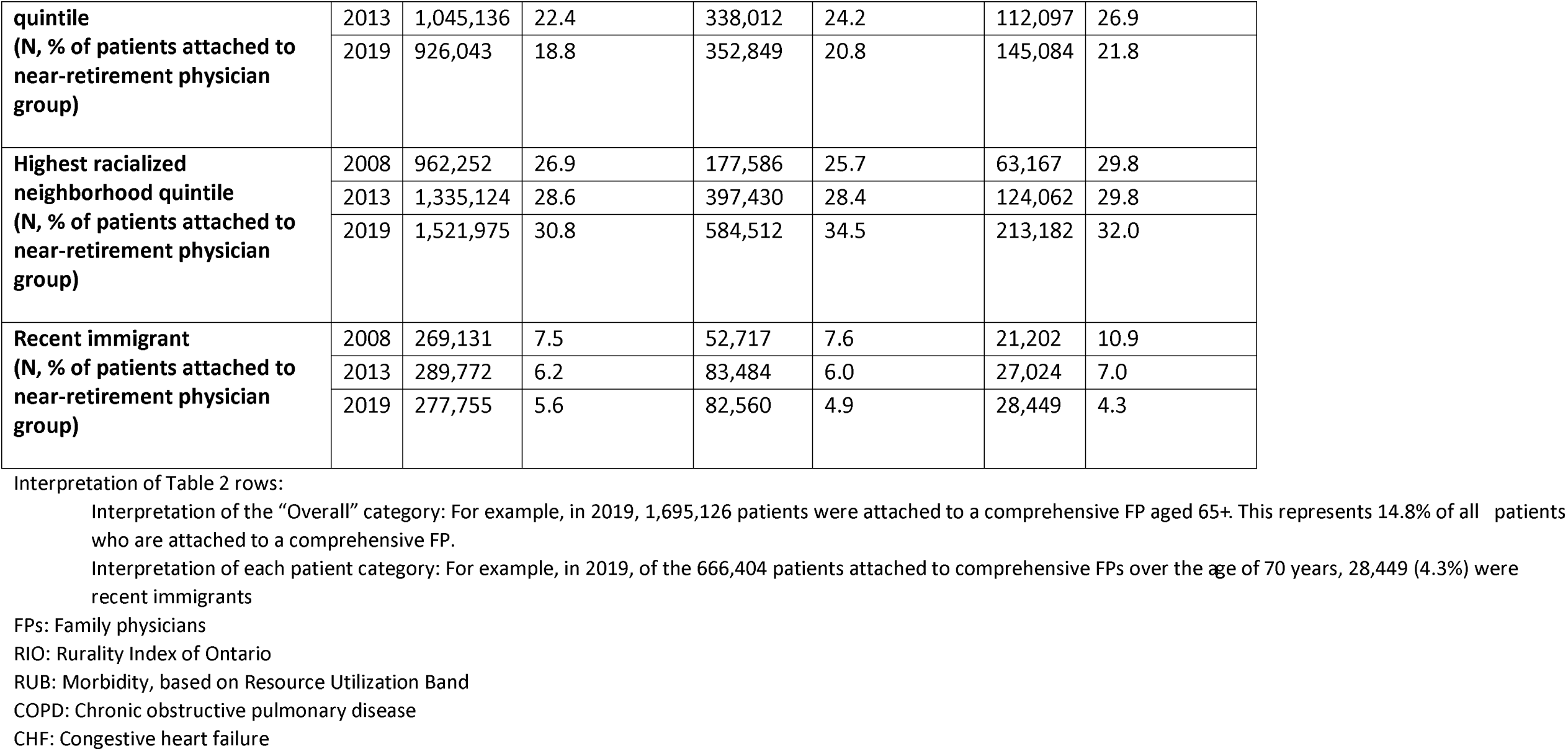
Characteristics of patients attached to near-retirement comprehensive family physicians over time, by near-retirement group

### Temporal Characteristics of Comprehensive Family Physicians and their Patients

#### Comprehensive FP Capacity/Workload

Table 1 shows the mean (SD) roster size for the total population of comprehensive FPs remained consistent over time (2008: 1213 (927); 2013: 1272 (909); 2019: 1209 (837)). Male FPs had consistently larger roster sizes in each age group and at each time point. Both male and female FP roster sizes followed an inverted U pattern with FP age, with practice sizes starting and ending smaller at the extremes of FP age and peaking during mid-career. This pattern was observed at all three time points with older (65+) male and female physicians and younger (<35) male and female physicians caring for larger roster sizes over time.

Working full time equivalent (FTE) also followed an inverted U pattern according to FP age (Table 1). Older physicians increasingly practiced FTE (2008: 58.4%, 2013: 67.0%, 2019: 72.6%). This was driven by an increasing proportion of female FTE comprehensive FPs. Among younger physicians, by 2019, females comprised the majority of FTE workforce (52.2% of FTE comprehensive FPs <35 years; 55.2% of FTE comprehensive FPs 35-44 years).

Mean (SD) annual core primary care visits provided per patient declined over time (Table 1): 2008: 7.3 (3.1) visits; 2013: 6.5 (2.6) visits; 2019: 6.0 (2.3) visits. In most comprehensive FP age groups, male and females provided similar numbers of annual visits. Older physicians provided more annual visits compared with their younger counterparts.

In the patient cohort (Table 2), at all near-retirement physician cut-offs (55+, 65+, 70+), a declining proportion over time made a high number (5+) primary care visits in the preceding year, but these proportions remained consistently over 50% in all near-retirement groups and at each time point.

#### Comprehensive FP Practice Settings

A declining proportion of comprehensive FPs over time practiced in fee-for-service (FFS) models of care. Alternate payment plan models (APPs), specifically capitation/team-based models of care, were an increasingly common setting over time (Supplement: eFigure 2). In these APP models, physician compensation is primarily a lump sum payment per attached patient, with or without additional government funding for interdisciplinary health professional supports. In 2008, most comprehensive FPs worked in FFS-based models (76.6%), but by 2019, most practiced in APPs (55.4%). This shift was seen across all comprehensive FP age groups (Supplement: eTable 2). Correspondingly, an increasing proportion of patients were served in APP models: 2008: 26.5% (n = 2,526,116); 2013: 54.3% (n = 5,643,862); 2019: 61.5% (n = 7,064,109).

Over time, a stable majority of comprehensive FPs practiced in large urban and urban settings (Supplement: eTable 3A). After a decline in 2013, an increasing proportion and number practiced in rural/remote areas by 2019, but numbers did not return to 2008 levels (2008: 6.7%, n = 513; 2013: 5.1%, n = 410; 2019: 5.3%, 492). Trends around age and sex of rural comprehensive FPs resembled trends seen in the overall comprehensive FP population (Supplement: eTables 3B, 3C).

#### Patient complexity

The mean age (SD) of comprehensive FPs’ patients increased over time (Table 1): 2008: 33.5 (13.2) years; 2013: 36.5 (12.1) years; 2019: 38.1 (12.0) years. When stratified by physician age and sex, each physician age group served increasingly older patients. Male physicians cared for slightly older patients than did women in each physician age group and at each time point.

The number and proportion of patients aged 65 and older increased over time in each near-retirement group (Table 2). This number nearly quadrupled in the oldest (70+ years) FP group (2008: N = 45,414, 2019: N = 176,473).

Comprehensive FPs cared for a stable mean (SD) proportion of female patients over time (Table 1) (2008:53.2% (12.9); 2013: 53.1% (12.5); 2019: 52.9% (12.0). Female comprehensive FPs had a greater proportion of female patients than male physicians at all time points and in all age groups. The overall proportion of female patients was higher in younger physician age groups at all time points, equalizing as physicians aged.

When examining the patient cohort by near-retirement physician age groups, the proportion of female patients also remained stable at each time point (Table 2), with slightly lower proportions of female patients in the oldest near-retirement group.

Over time, an increasing proportion of comprehensive FPs’ practices were comprised of the highest morbidity patients (Resource Utilization Band (RUB) 4+): 2008: 16.5%; 2013: 18.1%; 2019: 19.8% (Table 3). When stratified by comprehensive FP age and sex, older male physicians cared for higher proportions of the highest morbidity patients than did older female physicians in 2008 (65-69 years) and 2013 (65-69 years, 70+ years), but by 2019, males and females cared for similar proportions of highest morbidity patients within each and across all physician age groups.

**Table 3.** Practice characteristics: Medical and social complexity of patients attached to comprehensive family physicians over time by physician age and sex

|  |  | <35 Years |  |  | 35-44 Years |  |  | 45-54 Years |  |  | 55-64 Years |  |  | 65-69 Years |  |  | 70+ Years |  |  | TOTAL |  |  |
| --- | --- | --- | --- | --- | --- | --- | --- | --- | --- | --- | --- | --- | --- | --- | --- | --- | --- | --- | --- | --- | --- | --- |
|  |  | Total | M | F | Total | M | F | Total | M | F | Total | M | F | Total | M | F | Total | M | F | Total | M | F |
|  |  | % | % | % | % | % | % | % | % | % | % | % | % | % | % | % | % | % | % | % | % | % |
| **Highest morbidity (RUB 4+)** | 2008 | 15.3 | 14.7 | 15.6 | 16.2 | 15.8 | 16.7 | 16.4 | 16.5 | 16.2 | 17.3 | 17.5 | 16.6 | 16.8 | 17.2 | 14.0 | 14.0 | 14.1 | 13.0 | 16.5 | 16.7 | 16.3 |
|  | 2013 | 17.5 | 17.6 | 17.4 | 18.2 | 17.5 | 18.7 | 17.7 | 17.8 | 17.6 | 18.1 | 18.5 | 17.3 | 19.5 | 20.0 | 17.5 | 20.1 | 20.5 | 17.9 | 18.1 | 18.3 | 17.8 |
|  | 2019 | 19.3 | 19.4 | 19.2 | 20.6 | 20.2 | 20.8 | 19.4 | 19.4 | 19.4 | 19.5 | 20.2 | 18.7 | 20.3 | 20.4 | 20.1 | 21.4 | 21.5 | 21.3 | 19.8 | 19.9 | 19.7 |
| **Lowest income quintile** | 2008 | 18.5 | 19.2 | 18.1 | 18.1 | 19.6 | 16.6 | 18.4 | 19.8 | 16.4 | 19.9 | 20.2 | 18.8 | 22.6 | 22.5 | 23.6 | 23.9 | 20.1 | 17.2 | 19.0 | 20.1 | 17.2 |
|  | 2013 | 18.9 | 20.6 | 18.0 | 17.2 | 19.1 | 16.0 | 18.0 | 19.4 | 16.4 | 18.4 | 19.5 | 16.5 | 20.5 | 20.4 | 21.2 | 24.0 | 24.2 | 22.5 | 18.3 | 19.6 | 16.7 |
|  | 2019 | 20.4 | 21.9 | 20.7 | 18.8 | 20.7 | 17.6 | 18.3 | 20.5 | 16.5 | 18.8 | 20.4 | 16.8 | 19.9 | 20.7 | 17.9 | 22.1 | 22.2 | 21.4 | 19.0 | 20.7 | 17.5 |
| **Highest housing instability quintile** | 2008 | 24.5 | 22.8 | 25.5 | 20.6 | 20.7 | 20.4 | 20.4 | 20.4 | 20.4 | 21.9 | 21.6 | 23.0 | 24.0 | 23.1 | 29.2 | 25.5 | 25.6 | 24.2 | 21.4 | 21.2 | 21.7 |
|  | 2013 | 26.0 | 23.6 | 27.2 | 21.8 | 20.9 | 22.5 | 19.9 | 20.4 | 19.4 | 20.8 | 20.6 | 21.3 | 21.7 | 21.8 | 21.2 | 24.5 | 24.1 | 26.6 | 21.4 | 20.9 | 21.9 |
|  | 2019 | 26.5 | 25.3 | 27.2 | 24.5 | 24.7 | 24.5 | 21.1 | 21.8 | 20.4 | 21.4 | 21.5 | 21.3 | 22.6 | 21.7 | 24.9 | 25.5 | 25.2 | 27.1 | 23.0 | 22.7 | 23.3 |
| **Highest material deprivation quintile** | 2008 | 18.6 | 19.8 | 17.9 | 17.4 | 19.3 | 15.5 | 18.2 | 20.1 | 15.6 | 20.5 | 21.3 | 18.1 | 23.7 | 23.9 | 22.4 | 25.7 | 26.2 | 21.3 | 19.0 | 20.6 | 16.4 |
|  | 2013 | 22.9 | 24.6 | 22.0 | 20.5 | 22.1 | 19.4 | 21.2 | 22.9 | 19.3 | 21.4 | 22.6 | 19.2 | 23.7 | 23.2 | 25.7 | 29.2 | 29.4 | 27.8 | 21.5 | 22.8 | 19.9 |
|  | 2019 | 18.2 | 19.7 | 17.3 | 17.3 | 19.9 | 15.8 | 17.0 | 19.3 | 15.0 | 18.1 | 19.8 | 15.9 | 19.7 | 20.9 | 16.7 | 21.8 | 22.1 | 19.9 | 17.8 | 19.8 | 15.9 |
| **Highest racialized neighborhood quintile** | 2008 | 27.4 | 30.8 | 25.5 | 27.5 | 28.4 | 26.5 | 26.0 | 26.1 | 25.9 | 27.2 | 26.3 | 30.4 | 28.0 | 26.4 | 37.2 | 32.6 | 32.8 | 30.7 | 26.9 | 26.9 | 27.0 |
|  | 2013 | 29.9 | 31.1 | 29.2 | 28.6 | 29.2 | 28.2 | 27.9 | 29.2 | 26.6 | 27.2 | 27.2 | 27.3 | 27.7 | 25.5 | 37.3 | 33.0 | 32.0 | 39.4 | 28.0 | 28.1 | 28.0 |
|  | 2019 | 26.0 | 26.6 | 25.7 | 25.8 | 27.2 | 25.0 | 28.5 | 29.2 | 27.8 | 27.0 | 26.8 | 27.3 | 33.2 | 33.7 | 31.9 | 32.1 | 30.9 | 38.5 | 27.4 | 28.3 | 26.7 |

Interpretation: For example, in 2008, within the group of comprehensive family physicians under the age of 35 years, 15.3% of patients in those practices had the highest level of morbidity (RUB 4+). When further stratified by physician sex, 14.7% of patients attached to male comprehensive family physicians belonged to the highest morbidity (RUB 4+) group.
RUB: Morbidity, based on Resource Utilization Band

Table 2 shows the number and proportion of highest morbidity patients attached to near-retirement physicians grew over time. By 2019, 983,818 patients in the highest morbidity patients were attached to a physician aged 55+, representing 19.9% of all patients attached to a 55+ physician. 350,439 were attached to a 65+ physician (20.7% of patients attached to a 65+ physician). 146,298 were attached to a 70+ physician (22.0% of patients attached to 70+ a physician), representing a tripling of the absolute number.

While proportions of patients with chronic illness (COPD, CHF, diabetes, frailty, mental illness) remained relatively stable over time, the absolute numbers increased markedly in each near-retirement group (Table 2).

The proportions and means of socially complex patients cared for within each comprehensive FP age and sex group increased over time for most indicators (Table 3) and the number of higher social complexity patients increased markedly over time for most near-retirement groups (Table 2).

## DISCUSSION

In our population-level serial cross-sectional analyses, the proportion of patients attached to a comprehensive FP in Ontario, Canada, grew over time. However, we found an increasing proportion of the comprehensive FP workforce is nearing retirement. Given the average FP retires at age 70.5 years,^38^ we anticipate that by 2025, nearly 1.7 million Ontarians may lose their comprehensive FP to retirement, eroding gains made to date.

This number may be an underestimate for several reasons. First, half of all comprehensive FPs are now female, and female FPs retire on average 5 years earlier than males.^38^ Second, a decreasing proportion of FPs are practicing comprehensive family medicine. This trend was seen across every physician age group, indicating practicing FPs are leaving comprehensive primary care earlier in their careers than in previous years while a smaller proportion of incoming FPs are choosing to enter comprehensive practice. Third, due to limitations in data availability for more recent years, our analyses predate the COVID-19 pandemic, and surveys from Ontario indicate the pandemic has hastened retirement plans, with almost double the usual proportion of FPs closing their offices during the pandemic (3%, compared with the usual rate of 1.6%/year),^41^ and one in five indicating an intention to retire within five years.^42^

Several other trends identified likely apply to other jurisdictions nationally and internationally and, when taken together, indicated limited capacity in the workforce to absorb the workload of near-retirement physicians. Comprehensive FPs cared for increasingly older groups of patients with increasing complexity over time. As of 2019, all physician age groups served similar proportions of complex patients, and near-retirement physicians cared for an increasing number and proportion of older patients with increasing medical and social complexities. Females, who comprised an increasing proportion of the comprehensive FP workforce, served similar proportions of highest morbidity patients but smaller roster sizes compared with males, which may reflect previous research finding women primary care physicians spend more time with and receive more requests from patients.^31 32^ That said, both the oldest and youngest male and female comprehensive FP groups served increasingly larger rosters, and an increasing proportion of older (65+) physicians practiced FTE.

Ontario continues to add a net positive number of FPs to the workforce each year, but this number has declined from 453 in 2017 to 303 in 2020.^43^ Over the past 7 years, a smaller proportion of medical school graduates ranked family medicine as their first choice discipline,^44^ echoing trends in other jurisdictions including the United Kingdom and the United States.^20-22^ The future supply of incoming FPs may therefore be inadequate to meet needs identified in our study, especially considering the 1.6 million Ontarians already without a regular primary care provider in our 2019 cohort.

Solutions to FP workforce shortages identified in the literature focus on addressing deterrents to the practice of comprehensive primary care, including perceived poor respect for primary care as a profession, inadequate compensation, inadequate training supports for developing and maintaining comprehensive skills, and inadequate administrative and interdisciplinary health supports to manage increasing patient complexity.^21 24 45-49^ Our finding of a shift toward APP models underscores the desire among comprehensive FPs for financial stability and team-based supports. Further, we identified large numbers of patients with chronic diseases and complex social needs, all of which are highly amenable to team-based care.^50-52^

There are some limitations to our study. The FTE indicator is based on physician billings and excluded non-billable administrative time. Almost half of Canadian FPs report 10-19 hours per week of administrative tasks,^53^ so the indicator may underestimate workload, and thus the number of FTE FPs. Rural FPs often practice in both primary care and hospital settings;^54^ since the comprehensiveness algorithm is based on primary care billings,^29^ it may underestimate the number of rural comprehensive FPs. Further, the rurality index scores and methodology have not been updated since 2008. Some physician analyses could not be fully stratified by both age and sex due to small cell sizes. Community Health Centre patients are not included and we did not examine other clinicians who may provide primary care; however, these clinicians are the main primary care source for only a small minority of Ontarians.^55 56^ Finally, our analyses do not account for the rise of virtual care and its potential impact on capacity.^57-59^

## CONCLUSIONS

Primary care faces many capacity challenges as physicians age into retirement and fewer choose to enter or remain in comprehensive practice. Incentives and supports are needed to grow the comprehensive FP workforce to serve a growing and increasingly complex patient population.

## Supporting information

Supplemental eFigure 1. Proportion of family physicians practicing comprehensiveness by year, age, and sex

Supplemental eFigure 2. Proportion of comprehensive family physicians in various practice models by year

## Data Availability

The data sets from this study are held securely in coded form at ICES. Data-sharing agreements prohibit ICES from making the data sets publicly available, but access may be granted to those who meet pre-specified criteria for confidential access, available at www.ices.on.ca/DAS. The complete data set creation plan, and underlying analytic code are available from the authors upon request, understanding that the programs may rely upon coding templates or macros unique to ICES.

## Author Contributions

*Concept and design*: Premji, Green, Glazier, Ryan

*Acquisition, analysis, or interpretation of data:* All authors

*Drafting of the manuscript:* Premji

*Critical revision of the manuscript for important intellectual content*: All authors

*Statistical analysis:* Premji, Khan, Ryan, Green, Glazier

*Obtained funding:* Green, Glazier

*Administrative, technical, or material support:* Frymire, Khan

*Supervision*: Ryan, Mathews

## Conflict of Interest Disclosures

None declared.

## Funding/Support

This study was supported by the INSPIRE PHC (Innovations Strengthening Primary Health Care Through Research) Research Program, which is funded through the Health Systems Research Program of the Ontario Ministry of Health (MOH) and the Ontario Ministry of Long-term Care (MLTC). It was also supported by ICES, which is funded by an annual grant from the Ontario MOH and MLTC. Dr. Premji was also supported by the PhD Family Medicine program at the University of Western Ontario, and by the Junior Clinical Research Chair in Family Medicine at the Department of Family Medicine, University of Ottawa.

## Role of the Funder/Sponsor

The funder had no role in the design and conduct of the study; collection, management, analysis, and interpretation of the data; preparation, review, or approval of the manuscript; and decision to submit the manuscript for publication.

## Disclaimer

The analyses, conclusions, opinions, and statements expressed herein are solely those of the authors and do not reflect those of the funding or data sources; no endorsement is intended or should be inferred.

## Additional Information

Parts of this material are based on data and/or information compiled and provided by CIHI and Cancer Care Ontario (CCO). The analyses, conclusions, opinions and statements expressed herein are solely those of the authors and do not reflect those of the data sources; no endorsement is intended or should be inferred.

## Notes

### Competing Interest Statement

The authors have declared no competing interest.

### Author Declarations

The use of data in this study was authorized under section 45 of Ontario's Personal Health Information Protection Act (PHIPA) and did not require review by a research ethics board or informed consent.

### Summary of Updates

1. Updated Background section to clarify the definition and roles of primary care. 2. Minor edits throughout for clarity and flow. 3. Wording change to better define the label of one covariate in Tables 2 and 3 (previously "Neighborhood Ethnic Concentration", now "Racialized Neighbourhood").

