## Supplemental Methods for "Trends in patient attachment to an aging primary care workforce: a population-based serial cross-sectional study in Ontario, Canada"

**eMethods. Data sources, cohort definitions, and variable definitions**

We obtained study data from population-level, de-identified, linked health administrative databases housed at ICES. ICES is an independent, non-profit research institute whose legal status under Ontario’s health information privacy law allows it to collect and analyze healthcare and demographic data, without consent, for health system evaluation and improvement. Secure access to these data is governed by policies and procedures that are approved by the Information and Privacy Commissioner of Ontario. In 2018, the institute formerly known as the Institute for Clinical Evaluative Sciences formally adopted the initialism ICES as its official name. This change acknowledges the growth and evolution of the organization’s research since its inception in 1992, while retaining the familiarity of the former acronym within the scientific community and beyond.

The dataset from this study is held securely in coded form at ICES. While legal data sharing agreements between ICES and data providers (e.g., healthcare organizations and government) prohibit ICES from making the dataset publicly available, access may be granted to those who meet pre-specified criteria for confidential access, available at [www.ices.on.ca/DAS](http://www.ices.on.ca/DAS) (email: ). The full dataset creation plan and underlying analytic code are available from the authors upon request, understanding that the computer programs may rely upon coding templates or macros that are unique to ICES and are therefore either inaccessible or may require modification.

These datasets were linked using unique encoded identifiers and analyzed at ICES.

The index date for each covariate was the fiscal year-end for each time point: March 31, 2008, March 31, 2013, March 31, 2019.

Physician-level data came from the ICES Physician Database (age, sex, years in practice, practice specialty, practice type, full-time equivalence), the Primary Care Population database (geographic location, roster size, primary care model), and Ontario Health Insurance Plan (OHIP) billings (health services rendered). For physicians for whom birth month and date were missing, we imputed physician age based on birth year, with fiscal year end (March 31) as the index date. Physician gender is not available in ICES data, so physician sex was used instead, available as male and female.

Patient-level data came from the Registered Persons Database (age, sex, postal code, immigration status), the Client Agency Program Enrolment (CAPE) database (primary care enrolment model), the Community Health Centre database (CHC) (patients receiving health services at CHCs, which serve vulnerable patients), census data holdings (income quintiles and other marginalization indices), OHIP database (health services claims and associated diagnoses), Discharge Abstract Database linkages with OHIP (mental health diagnosis), and Johns Hopkins Adjusted Clinical Groups (frailty, resource utilization band).

Resource Utilization Bands (RUB): This was measured using the Johns Hopkins Adjusted Clinical Groups (ACG) Version 10.0. The RUB measure assesses expected health care use as a measure of patient complexity/morbidity.

Annual number of core primary care visits were based on activity billing codes for 22 primary care service types in the 12 months preceding the index date.

Rurality: We measured rurality using the practice postal code and the Rurality Index for Ontario (RIO) scoring methodology,^1^ with the following categories: Large urban (score 0), Urban (score 1-9), Small Urban/Suburban (score 10–39), and Rural/Remote (score ≥ 40).

Full-time equivalency (FTE): FTE was calculated based on payments from all sources, with a 40th percentile cut-point corresponding with a FTE of 1.0.

Chronic diseases (COPD, CHF, Diabetes): These were measured using validated cohorts at ICES. The algorithm used to define cohorts varies slightly for each chronic condition, based on the original ICES algorithm for diabetes (i.e., two physician claims or one hospital admission with diabetes within two years). These disease cohorts are cumulative over time.

Frailty: This was measured using the Johns Hopkins Adjusted Clinical Groups (ACG) Version 10.0 frailty defining diagnoses indicator, which captures patients with multidimensional frailty at the population level and is based on 10 clusters of frailty defining dimensions: Malnutrition, dementia, impaired vision, decubitus ulcer, incontinence of urine, loss of weight, poverty, barriers to access to care, difficulty in walking, and falls. The indicator has been demonstrated to accurately identify patients with limitations in activities of daily living.

Mental illness: The case definition algorithm to identify patients with a mental health diagnosis over the last two years links two databases at ICES: The Discharge Abstract Databasae (DAD) and OHIP. It is based on having two physician billing claims in OHIP over 2 years or one hospitalization with one of the listed mental health service codes (ICD9/ICD10).

Marginalization: We assessed three dimensions of marginalization (residential instability, material deprivation, and neighborhood ethnic concentration (termed “racialized neighborhood” in our tables)) using the Ontario Marginalization Index,^2^ a census-derived geographically-based index.

Physician-level continuity of care (CoC): The algorithm considers patients to be virtually attached a primary care physician if they received the majority of their primary care over the preceding 2-year period from a physician with greater than 10% physician-level continuity of care (CoC). Physician-level CoC is a visit-based measure of the proportion of an individual physician visits over all physician’s visits over a two-year time period. The numerator is the number of patients virtually attached to a physician, and the denominator is all unique patients the same physician had seen over two years. If the physician CoC is less than or equal to 10%, then this physician had a low CoC.
