## Supplemental eFigure 1. Proportion of family physicians practicing comprehensiveness by year, age, and sex for "Trends in patient attachment to an aging primary care workforce: a population-based serial cross-sectional study in Ontario, Canada"

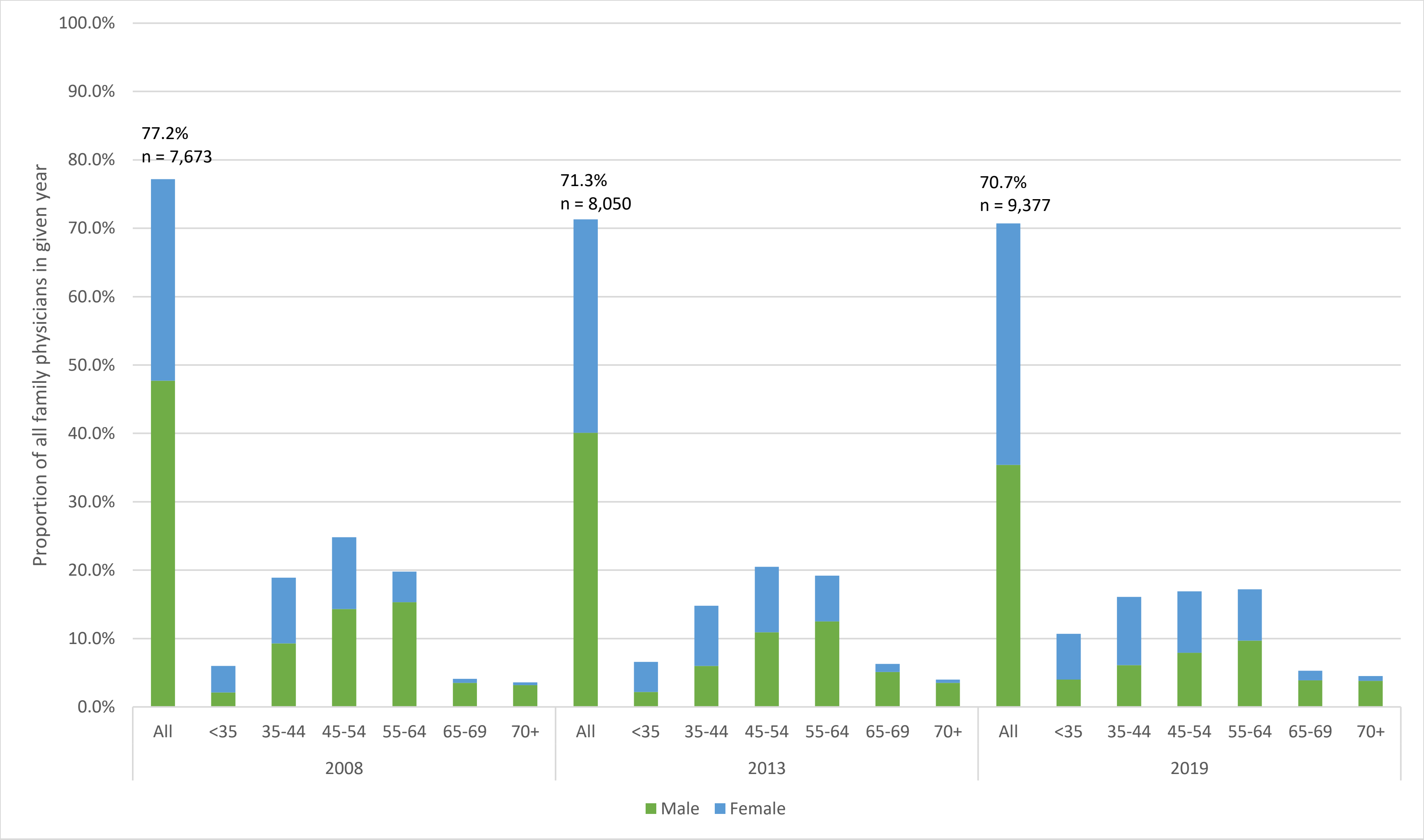


Total Ns (all family physicians): 2008: 9,944; 2013: 11,288; 2019: 13,269
