## Supplemental eFigure 2. Proportion of comprehensive family physicians in various practice models by year for "Trends in patient attachment to an aging primary care workforce: a population-based serial cross-sectional study in Ontario, Canada"

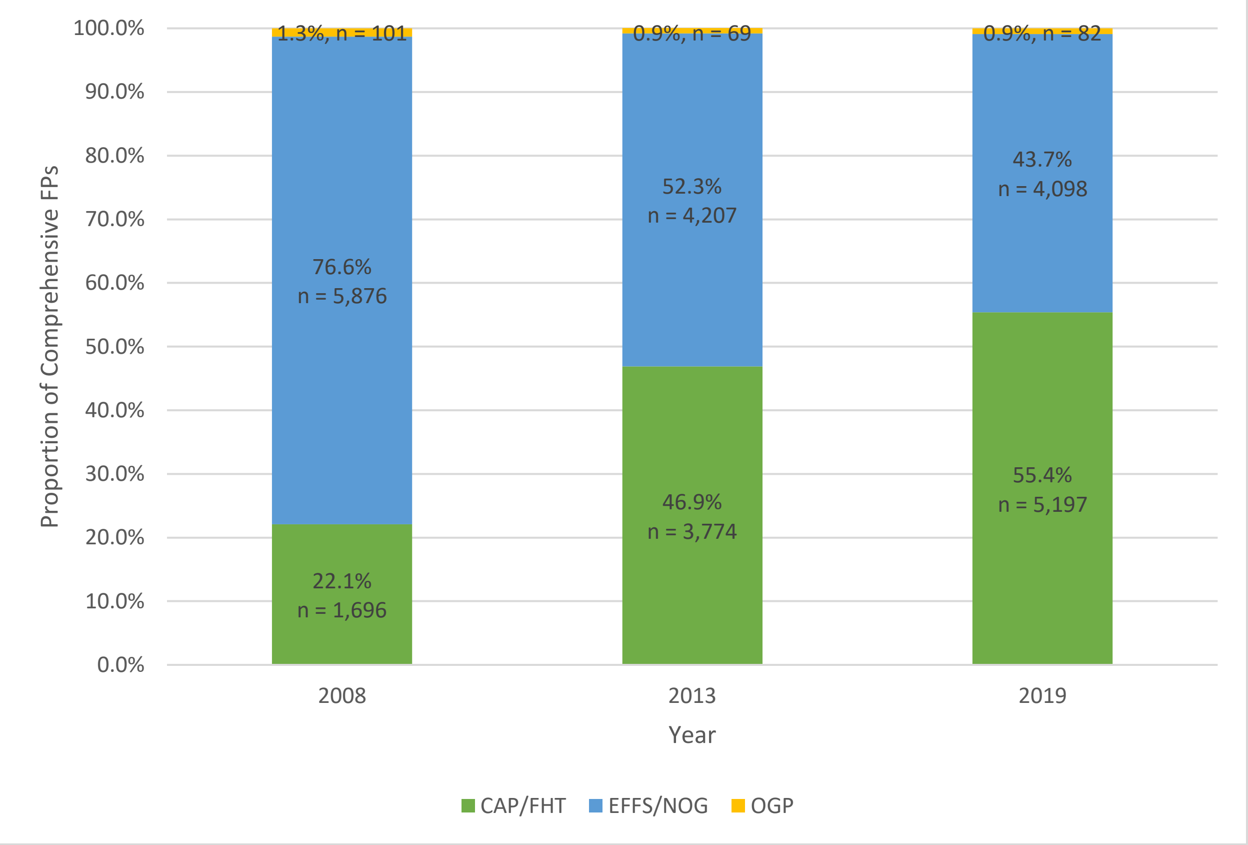


Total Ns (all comprehensive family physicians): 2008: 7,673; 2013: 8,050; 2019: 9,377

CAP/FHT: Alternate payment plan (APP) model where physician payments are mainly capitation(CAP)-based (annual amount per enrolled patient, adjusted for patient age and sex), with or without additional funding for interdisciplinary team members (Family Health Team(FHT)) such as nurse practitioners and social workers
EFFS/NOG: Fee-for-service payment models. EFFS = fee-for-service payments with enrolment requirements and some pay enhancements, such as higher payments for enrolled patients and bonus payments for meeting preventive care targets. NOG = No group; traditional fee-for-service payments with no enrolment requirements or payment enhancements.
